# Prevalence, Severity and Mortality associated with COPD and Smoking in patients with COVID-19: A Rapid Systematic Review and Meta-Analysis

**DOI:** 10.1101/2020.03.25.20043745

**Authors:** Jaber S. Alqahtani, Tope Oyelade, Abdulelah M. Aldhahir, Saeed M. Alghamdi, Mater Almehmadi, Abdullah S Alqahtani, Shumonta Quaderi, Swapna Mandal, John R. Hurst

## Abstract

**Background:** Coronavirus disease 2019 (COVID-19) is an evolving infectious disease that dramatically spread all over the world in the early part of 2020. No studies have yet summarised the potential severity and mortality risks caused by COVID-19 in patients with chronic obstructive pulmonary disease (COPD), and we update information in smokers.

**Methods:** We systematically searched electronic databases from inception to March 24, 2020. Data were extracted by two independent authors in accordance with the Preferred Reporting Items for Systematic Reviews and Meta-Analyses guidelines. Study quality was assessed using a modified version of the Newcastle-Ottawa Scale. We synthesised a narrative from eligible studies and conducted a meta-analysis using a random-effects model to calculate pooled prevalence rates and 95% confidence intervals (95%CI).

**Results:** In total, 123 abstracts were screened and 61 full-text manuscripts were reviewed. A total of 15 studies met the inclusion criteria, which included a total of 2473 confirmed COVID-19 patients. All studies were included in the meta-analysis. The crude case fatality rate of COVID-19 was 6.4%. The pooled prevalence rates of COPD patients and smokers in COVID-19 cases were 2% (95% CI, 1%–3%) and 9% (95% CI, 4%–14%) respectively. COPD patients were at a higher risk of more severe disease (risk of severity = 63%, (22/35) compared to patients without COPD 33.4% (409/1224) [calculated RR, 1.88 (95% CI, 1.4– 2.4)]. This was associated with higher mortality (60%). Our results showed that 22% (31/139) of current smokers and 46% (13/28) of ex-smokers had severe complications. The calculated RR showed that current smokers were 1.45 times more likely [95% CI: 1.03–2.04] to have severe complications compared to former and never smokers. Current smokers also had a higher mortality rate of 38.5%.

**Conclusion:** Although COPD prevalence in COVID-19 cases was low in current reports, COVID-19 infection was associated with substantial severity and mortality rates in COPD. Compared to former and never smokers, current smokers were at greater risk of severe complications and higher mortality rate. Effective preventive measures are required to reduce COVID-19 risk in COPD patients and current smokers.

## Introduction

The emergence in Wuhan City, Hubei Province of China of a novel pneumonia of unknown origin on the 31^st^ of December, 2019 was the start of an outbreak which would later be declared a pandemic by the World Health Organization (WHO) (1). The name COVID-19 (acronym for “coronavirus disease 2019”) was coined on the 11^th^ of February 2020 to describe presentation with severe acute respiratory disease (2). COVID-19 is caused by a novel strain of coronavirus, the severe acute respiratory syndrome coronavirus 2 (SARS-CoV-2). This virus belongs to the family of single stranded RNA viruses some of which have been previously described to be responsible for the Severe Acute Respiratory Syndrome (SARS) and Middle East Respiratory Syndrome (MERS) (3, 4). Although the symptoms and clinical presentation of COVID-19 is similar to SARS and MERS, the rate of spread is greater (5). As of 25^th^ of March 2020, the total number of confirmed case of COVID-19 stands at 459,419 with 20,818 deaths globally and the number is projected to increase (6).

Chronic Obstructive Pulmonary Disease (COPD) is a common, persistent and preventable dysfunction of the lung associated with limitation in airflow. COPD is a complex disease associated with abnormalities of the airway and/ or alveoli which is predominantly caused by exposure to noxious gases and particulates over a long period (7). With a global prevalence of 251 million cases in 2016, and 3.17 million (5%) deaths in 2015 alone, COPD was ranked by the WHO as the third leading cause of death especially with a particular burden in low- and middle-income countries (8, 9). This burden is predicted to grow due mainly to increased global exposure to tobacco, aging populations, poor awareness and inadequate access to diagnosis (10). COPD exacerbations are a major event in the natural history of the disease associated with worsening of symptoms often resulting in hospitalisation and poor prognosis (11). Various factors have been described to contribute to acute worsening of COPD, however, viral infection remains the main trigger, including seasonal coronaviruses (12-14).

Since the emergence of COVID-19, scientists around the world continue to better understand the clinical, diagnostic and prognostic characteristic of the disease. While over 150 papers, editorials and comments have been written about COVID-19 as of 24^th^ of March 2020, there is none dedicated to the specific risk posed to patients with a previous history of COPD. This review addresses this gap in knowledge with the aim of assisting clinicians to assess the prognosis of COVID-19 infection in patients with COPD and those with smoking history.

## Methods

This systematic review was conducted in accordance with the Preferred Reporting in Systematic Reviews and Meta-Analyses (PRISMA) guidelines (15). We prospectively submitted this review to Prospero (ID: 175518).

We searched MEDLINE and Google scholar from inception date to March 16, 2020. The search was updated on March 24, 2020. We used an extensive search strategy developed by a specilaised librarian for retrieving this type of evidence, which included the reference list of eligible papers and published pre-print papers (see supplementary Table S1). All retrieved studies were exported into EndNote to remove duplicates. The remaining studies were exported to Rayyan software for title, abstracts and full text screening by two independent reviewers.

### Inclusion and exclusion Criteria

Eligible studies were those that investigated: epidiomolgical, clinical charcterstics and features of COVID-19 and prevelance of chronic diseases specifically COPD in their analysis. We excluded the following: studies that did report COPD, studies that included respiratory diseases but did not specifically analyse COPD, only children, editorials, correspondence letters, reviews, qualitative studies, theses, non-English manuscripts and non-full text articles.

### Data Collection

Two authors independently screened titles and abstracts of potential studies and conflicts were resolved through discussion between the two. Full-text articles of potential studies were then independently read by two authors to identify studies meeting the inclusion criteria. The reference lists from all identified studies and reviews were scrutinized for eligible articles. Disagreement on selected papers was resolved through discussion with a third author.

### Quality Assessment

Two authers independently evaluated the methodological quality of included studies using a modified version of the Newcastle-Ottawa Scale (NOS) (16). It includes seven domains, each one of these domains was scored from 0 (high risk of bias) to 3 (low risk of bias) and we took a mean of the domains to result in a score between 0 and 3, where a higher score represents a lower risk of bias. Any disagreement in the quality assessment was resolved by discussion with a third author.

### Data Synthesis

We completed meta-analysis to calculate the pooled prevalence of COPD and current smokers among those patients with confrmed COVID-19. The output was generated using the Stata procedure Metaprop. Owing to heterogeneity within and between studies, we used the random-effects model in Stata/SE 15. Data were displayed using forest plots. We examined between-study heterogeneity using the I^2^ statistic. A narrative synthesis of the results was conducted considering the prevelance, disease severity and mortality among COVID-19 COPD patients and smoking status. We defined COVID-19 severity as those who were admitted to intensive care unit (ICU), had severe, oxygenation, needed mechanical ventilation or death.

### Pre-Print

As this is a rapidly evolving area, we also searched pre-print literature and whilst not incorporating such data in the formal analysis, provide narrative synthesis where such pre-print data are relevant to our main findings.

## Results

An initial search generated 123 potentially relevant papers, of which two were immediately excluded due to duplication. After the first screening of title and abstracts, 61 papers were potentially relevant according to the inclusion criteria. An additional 46 papers were excluded after full-text review, which resulted in 15 studies that satisfied all criteria. The reference list of the relevant papers was also examined (see Figure 1, PRISMA flow diagram).

**Figure 1.**
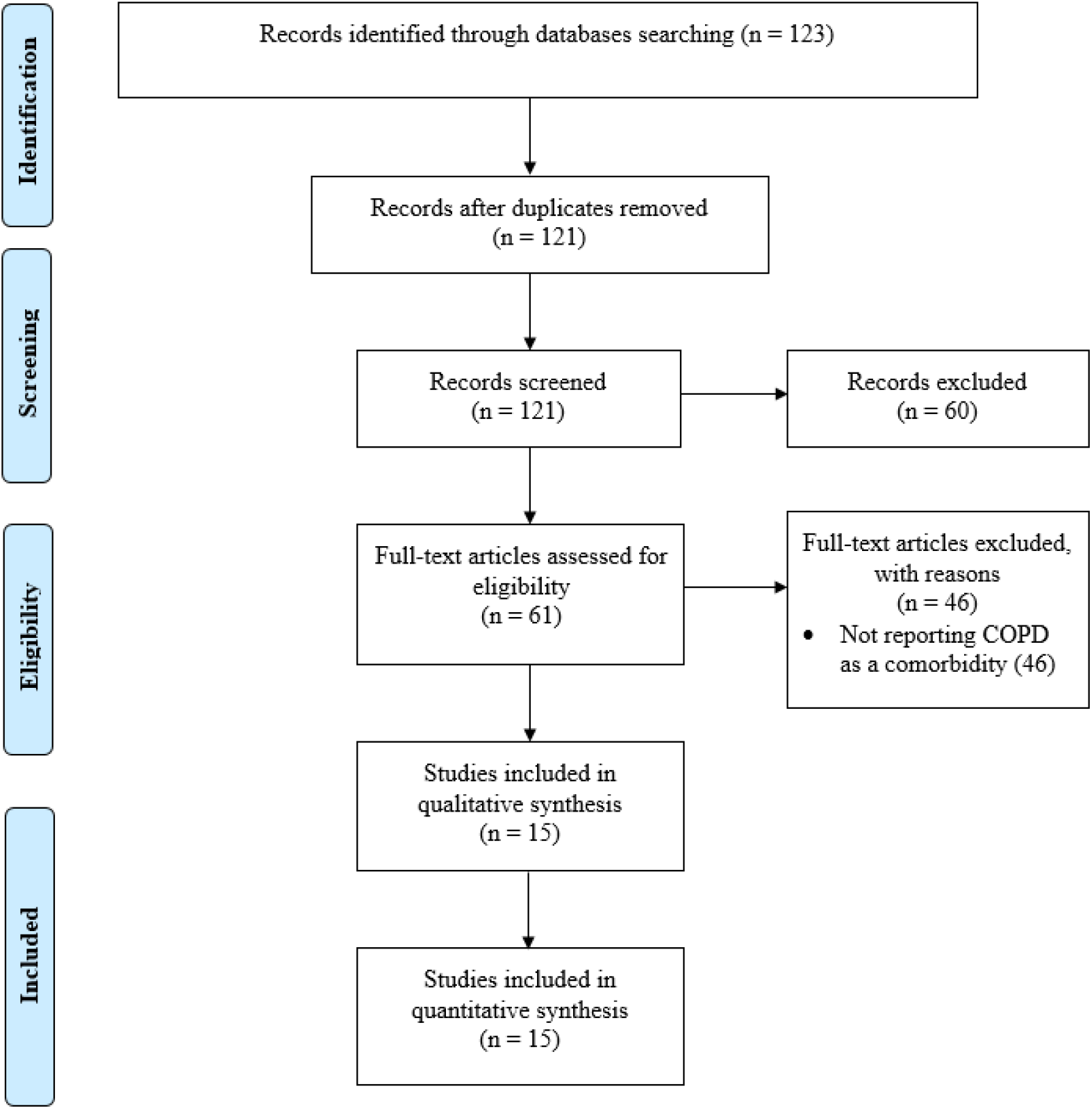
PRISMA flow chart for the included studies.

### Description of included studies

A summary of the included studies is presented in Table 1, which included a total of 2473 confirmed COVID-19 patients. Of those patients, only 58 (2.3%) had COPD as a comorbidity. The sample size of the included studies ranged from 21 to 1099 patients. Most of the studies were retrospectively conducted and all were studied in China, except one in the United States. The risk of bias ranged from 0.4 to 2.7; nine studies scored ≥ 2, which indicates low risk of bias (see, Table S2). The overall crude mortality rate for COVID-19 was 7.4% (184/2473). There were no reports of the baseline characteristics of the 58 COPD patients that described their age, or severity of COPD airflow limitation. Smoking status was reported in eight studies, in which 221 confirmed COVID-19 cases were current smokers.

**Table 1.** Characteristics of the included studies.

| Study Name | Country/ Type of study | Sample size<br>(male,<br>Female) | Age<br>mean ±SD<br>median or<br>range | Current<br>Smokers | COPD<br>patients | COPD<br>Survived | COPD<br>Non-Survived | Mortality<br>rate for<br>study/COPD | Severe cases for<br>study/ severe<br>COPD Cases |
| --- | --- | --- | --- | --- | --- | --- | --- | --- | --- |
| Arentz M et al.<br>2020 (17) | United states/ Washington<br>Retrospective study from February<br>20, 2020, and March 5, 2020, | 21<br>M= 11 (52%)<br>F= 10 (48%) | 70 (43-59) | NA | 7<br>(33.3%) | NA | NA | 11 (52.4%) | NA |
| Guan et al. 2020<br>(18) | China/ 30 provinces<br>Retrospective study from December<br>11, 2019, and January 29, 2020 | 1099<br>M= 640 (58.3%)<br>F= 459 (41.7%) | 47 (35–58)<br>Severe<br>52 (40–65)<br>Non-severe<br>45 (34–57) | 137 (12.5%)<br>Severe cases=<br>29/172<br>(16.9%) | 12<br>(1.1%) | NA | NA | 15 (1.4%) | 173 (15.7%)/ 6 (3.4%) |
| Huang Y et al.<br>2020 (19) | China /Hubei province<br>Retrospective study from December<br>2019 to January 2020 | 34<br>M= 14 (41.2%)<br>F= 20 (58.8%) | 56.24 ±<br>17.14 | NA | 3<br>(8.82%) | NA | NA | NA | NA |
| Huang et al. 2020<br>(20) | China/Wuhan<br>Prospective study from Dec 16, 2019,<br>to Jan 2, 2020 | 41<br>M= 30 (73%)<br>F= 11 (27%) | 49 (41–58) | 3 (7%) | 1 (2%) | NA | NA | 6 (15%) | 29 (70.7%)/ 1 (3.44%) |
| Liu et al. 2020<br>(21) | China/Hubei province<br>Retrospective study from December<br>30, 2019 to January 24, 2020 | 137<br>M= 61 (44.5%)<br>F= 76 (55.5%) | 57 (20–83) | NA | 2 (1.5%) | NA | NA | 16 (11.7%) | NA |
| Mo et al. 2020<br>(22) | China/Wuhan<br>retrospective study from January 1st<br>to February 5th | 155<br>M= 86 (55.5%)<br>F= 69 (45.5%) | 54 (42-66) | 6 (3.9%) | 5 (3.2%) | NA | NA | NA | 85 (54.8%)/ 4 (3.7%) |
| Wang et al. 2020<br>(23) | China/Wuhan<br>Retrospective, single-center case<br>series from January 1 to January 28,<br>2020 | 138<br>M= 75 (54.3%)<br>F= 63 (45.7%) | 56 (42-68)<br>ICU (n =<br>36)= 66 (57-78)<br>Non-ICU (n<br>= 102)= 51<br>(37-62) | NA | 4 (2.9%) | NA | NA | 6 (4.3%) | 3 (8.3%) |
| Wu et al. 2020 (24) | China/Wuhan<br>Retrospective study from December 25, 2019, and January 26, 2020 | 201<br>M=128 (63.7%)<br>F= 73 (36.3%) | 51 (43-60) | NA | 5 (2.5%) | NA | NA | 44 (21.9%) | NA |
| Wu J et al. 2020 (25) | China/ outside of the Wuhan area)<br>Prospective study from January to February 2020 | 80<br>M= 42 (52%)<br>F= 38 (48%) | 44 (11) | 26 (33%) | 3 (4%) | NA | NA | NA | NA |
| Xu X et al. 2020 (26) | China/Guangzhou<br>retrospective study from January 23, 2020, and February 4, 2020 | 90<br>M=39 (43%)<br>F= 51 (57%) | 50 (18–86) | NA | 1 (1%) | NA | NA | NA | NA |
| Xu XW et al. 2020 (27) | China/Zhejiang province<br>Retrospective study from 10 January 2020 to 26 January 2020 | 62<br>M= 35 (56%)<br>F= 27 (44%) | 41 (32-52) | NA | 1 (2%) | NA | NA | 0 | 45 (72.5%)/ 1 (3%) |
| Yang et al. 2020 (28) | China/Wuhan<br>Retrospective study from late December, 2019, and Jan 26, 2020. | 52<br>M= 35 (67%)<br>F= 17 (33%) | 59.7 (13.3) | 2 (4%)<br>Survived=2 (10%) | 4 (8%) | 2 (10%) | 2 (6%) | 32 (61.5%)/2 (50%) | NA |
| Zhang et al. 2020 (29) | China/Wuhan<br>Retrospective study from January 16 to February 3, 2020 | 140<br>M=71 (50.7%)<br>F= 69 (49.3%) | 57 (25-87) | 2 (1.4%)<br>Severe cases= 2 (3.4) | 2 (1.4) | NA | NA | NA | 58 (41.4%)/ 2 (3.4%) |
| Zhou et al. 2020 (30) | China/Wuhan<br>retrospective cohort study from Dec 29, 2019 to Jan 31, 2020 | 191<br>M= 119 (62%)<br>F= 72 (38%) | 56 (46–67) | 11 (6%)<br>Non-survivor= 5 (9%)<br>Survivor= 6 (4%) | 6 (3%) | 2 (1%) | 4 (7%) | 54 (28.3%)/4 (66.7%) | NA |
| Zhu et al. 2020 (31) | China/ Anhui province<br>Retrospective study from 24 January 2020 and 20 February 2020 | 32<br>M= 15 (47%)<br>F= 17 (53%) | 46(35-52) | 6 (18.7%) | 2 (6%) | NA | NA | NA | NA |

### Prevalence of COPD in confirmed COVID-19 cases

The pooled prevalence of COVID-19 patients who had COPD from all studies was 2% (95% CI, 1%–3%), Figure 2.

**Figure 2.**
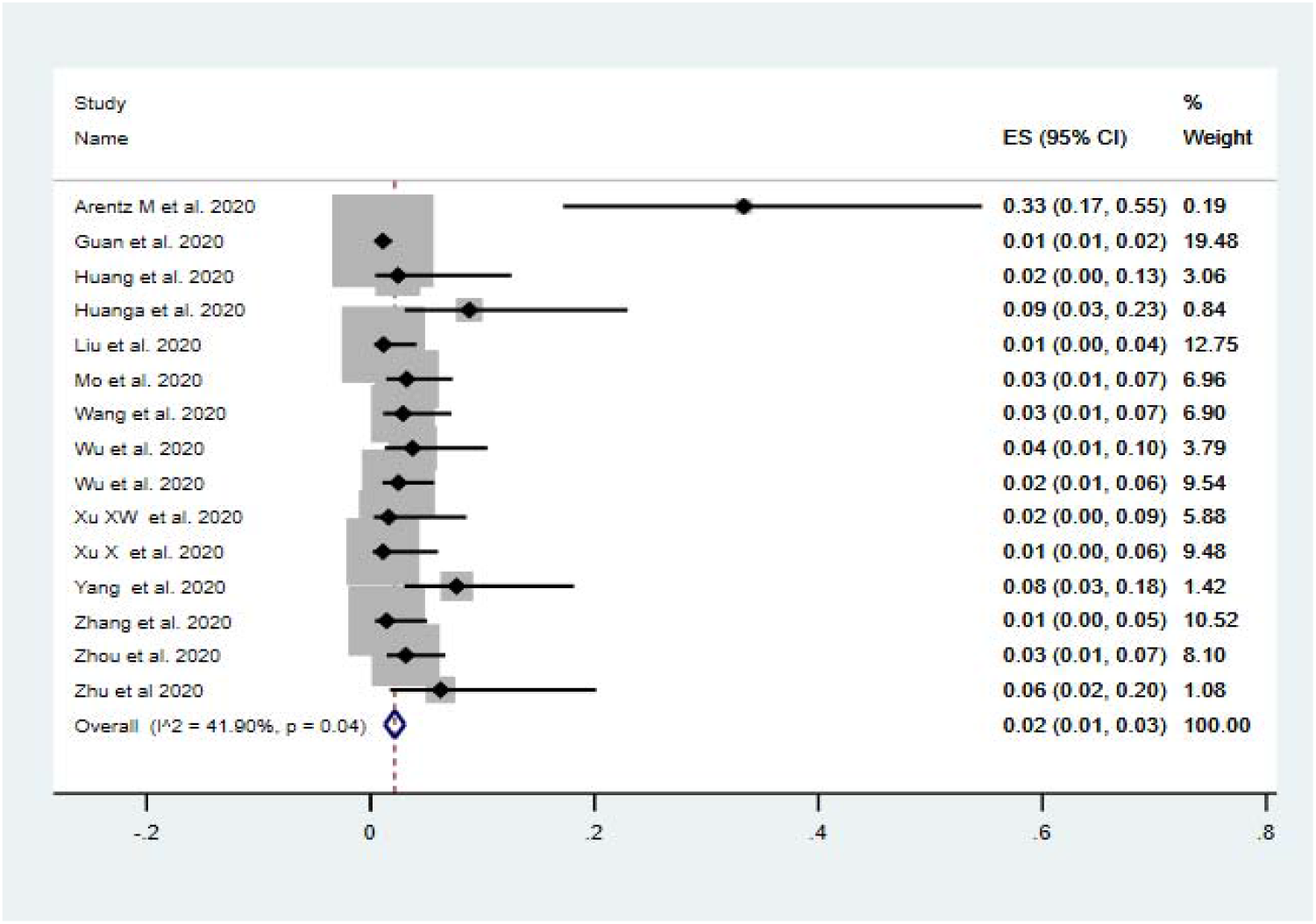
Pooled prevalence of COVID-19 patients with COPD.

### Disease severity and mortality among COVID-19 COPD patients

Seven studies that included 35 COPD patients reported COVID severity in their analysis. With 63% (22/35) patient reported as severe compared to 37% (13/35) non-severe, this shows that COPD patients are at a higher risk of more severe COVID-2019 compared to patients without COPD 33.4% (409/1224) [calculated RR, 1.88 (95% CI, 1.4–2.4)]. Data from two studies (28, 30) including COPD patients with confirmed COVID-19 demonstrate 60% (6/10) mortality rate compared to mortality rate in patients without COPD 55% (86/157), [calculated RR, 1.10 (95% CI, 0.6–1.8)].

### Smoking history and risk of COVID-2019

Smoking exposure including (current and ex-smokers) was reported in eight studies, with 221 confirmed COVID-19 cases. We assessed the prevalence of current smokers in all studies using meta-analysis of proportions of current smokers that had confirmed COVID-19. There was a pooled prevalence of 9%, (95% CI, 4%–14%), Figure 3. 22.30% (31/139) of current smokers and 46% (13/28) of ex-smokers had severe complications. Calculating the RR from these two studies showed that current smokers (31/108) were 1.45 times more likely [RR=1.45, 95% CI: 1.03–2.04] to have severe complications compared to former and never smokers (147/808) patients (18, 29). A higher mortality rate of 38.5% (5/13) was also seen among current smokers.

**Figure 3.**
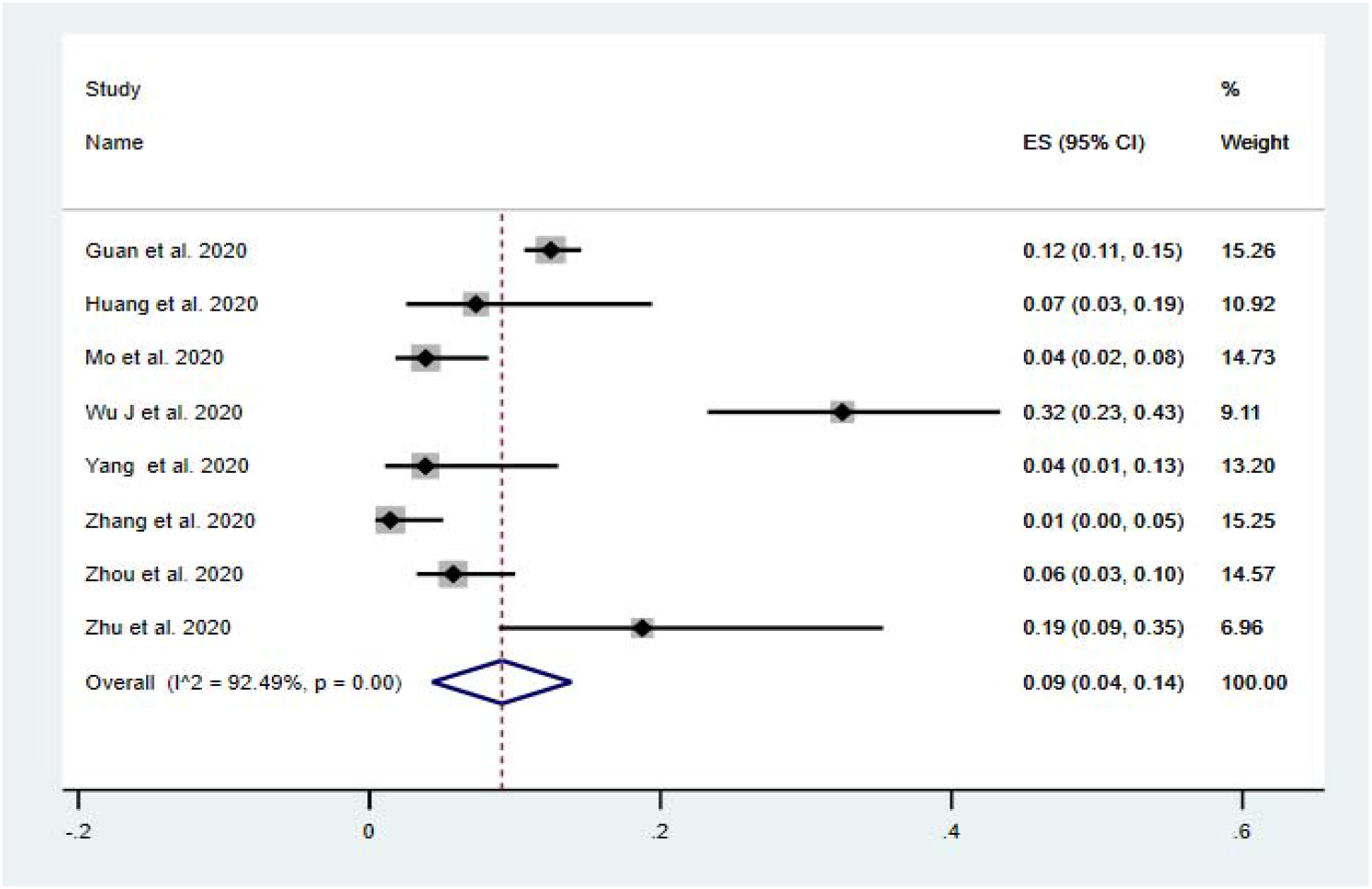
Pooled prevalence of current smokers using meta-analysis of proportion of current smokers that had confirmed COVID-19.

## Discussion

To the best of our knowledge, this is the first systematic review and meta-analysis to develop an informed understanding of the prevalence, severity and mortality of COPD patients diagnosed with COVID-19. We provide an updated report in relation to smokers (32). Our main outcomes show that the prevalence of COPD in COVID-19 patients was low, but that the risk of severity (63%) and mortality (60%) were high, which indicates COPD patients with confirmed COVID-19 are at a greater risk of severe complications and death. Furthermore, the prevalence of current smokers in COVID-19 patients was 9% (95% CI, 4%– 14%), and this was also associated with greater severity (22.30%) and mortality (38.5%).

We report a low prevalence of COPD patients in COVID-19 case series compared to the latest COPD prevalence rate in China, which was 13.6% (95% CI 12·0-15·2) and the global prevalence of COPD (9-10%) (33, 34). We speculate that patients may have not been diagnosed. Having a reliable estimate of the prevalence of COPD in COVID-19 cases, and likely outcomes, is crucial to ensure specific successful global preventive and treatment strategies for COPD patients. Bearing this in mind, in the included studies there was no report on COPD severity data and COPD-related comorbidities, which prevents us from assessing the impact of such essential information.

Although the COPD prevalence was not high in the included confirmed COVID-19 cases, COVID-19 causes a substantial burden on COPD patients with increased disease severity. This summary agrees with other results currently available only in pre-print (35, 36). Viral infections in COPD patients increase systemic inflammation with slow recovery of reported symptoms (37, 38). In addition to the effects of COVID-19, patients with COPD have various comorbidities, some of which are associated with increased risk of hospitalisation (39-41). According to recent systematic reviews, the prevalence of comorbidities in COVID-19 patients was high and these comorbidities were associated with increased disease severity (42, 43). Most of the studies that reported COPD severity defined severe cases as those who were admitted to intensive care unit (ICU), had severe oxygenation, needed mechanical ventilation or death (18, 19, 22, 23, 27, 29). These studies did not give details about the underlying COPD. It would have been interesting to know if the frequency of previous exacerbations was linked to increased complications in COPD patients diagnosed with COVID-19. In general, those with severe case of COVID-19 were older and had more coexisting comorbidities than those with mild illness.

Data from two studies (28, 30) describing COPD patients with confirmed COVID-19 show a higher mortality rate at 60%. This concurs with pre-print studies that found similarly high mortality rates (35, 44-47). Despite the small number of patients that were analysed, this increases concerns about the prognosis of this vulnerable population. However, this high mortality rate could be attributed to several factors. The majority of COPD patients have various comorbidities that may also be associated with mortality and related conditions may have been underreported because of the difficulties finding the specific cause of mortality (48). Moreover, in patients with severe COPD, respiratory failure is the principal cause of mortality and this demands ICU intervention. It is possible that limited access to respiratory support as part of COVID-19 management may be contributing to this mortality, dependent on critical care capacity in each hospital or region. According to a recent COVID-19 report from Italy, the surge in patients requiring intensive care has been unmanageable, with 12% of positive cases requiring ICU admission (49), more than that reported in China (18, 28). As a consequence, patients were dying because mechanical ventilation could not be offered, on top of acute shortage of clinicians who were able to manage those patients (50, 51). Until now, the particular mechanisms of how COVID-19 increases COPD severity and mortality is unknown, and undoubtedly more research is needed to find the possible mechanisms that linked COVD-19 and increased severity and mortality of COPD patients.

Concerning smoking and COVID-19, our data showed a pooled prevalence of 9% current smokers, (95% CI, 4%–14%), lower than the reported prevalence of smoking in China that was 25.2% (25.1–25.4) (52). Interestingly, we found that 22.30% (31/139) of current smokers and 46% (13/28) of ex-smokers had severe complications associated and greater mortality reaching 38.5% in current smokers. The calculated RR from two studies showed that current smokers were 1.45 times more likely [RR=1.45, 95% CI: 1.03–2.04] to have severe complications compared to former and never smokers (18, 29). The impact of smoking history on vulnerability to COVID-19 has been explored but there is limited data on the contribution of tobacco smoking to the spread of and poor outcome in COVID-19. A recent systematic review on COVID-19 and smoking including five studies found that smoking was most likely associated with the negative outcomes. This recommended further research to explore this in more detail due to limited studies (32). Evidence from other respiratory viruses, respiratory syncytial virus, has shown that inhaled tobacco smoke raises the transmission rate and severity of viral respiratory tract infections (53). It seems there is underlying mechanisms behind this prevalence, as smoking has been related to higher expression of ACE2 (the receptor for SARS-CoV-2) (pre-print) (54). However, as more reports globally from diverse racial and genetic contexts become available, differences in the production of ACE2 can be further evaluated and linked to how they lead to COVID-19 vulnerability in different groups (55).

To our knowledge, the present study is the first to systematically evaluate existing literature with a focus on risk of COVID-19 on COPD. For the first time, we conducted a meta-analysis using a random effects model to calculate the pooled prevalence of COPD in confirmed COVID-19 and examined outcomes. This increased the generalisability of our findings, as heterogeneity was addressed by incorporating between-study variability of effect sizes. We showed that COPD and smoking in COVID-19 is associated with greater disease severity and higher mortality. This review has some limitations. Few studies were eligible for inclusion and most of them come from China. Second, heterogeneity exists in location, setting, and design. The reported clinical characteristics were not available in most of the studies at the time of analyses. This work has a number of clinical and research implications. It highlights the global prevalence and the clinical effects of COVID-19 on COPD patients and smokers. As COPD patients are at an increased risk of severe outcomes if they became infected with COVID-19, it is recommended that patients and clinicians establish effective plans for ensuring prevention, such as using tele-medicine to ensure that COPD receive the best care (56, 57). We strongly advocate public awareness campaigns concentrating on ways to achieve smoking cessation among smokers, and it is possible that an improvement in cessation rates will help to reduce the spread of SARS-CoV-2. Future studies should investigate the mechanisms between COPD, smoking and COVID-19 infection.

## Conclusion

Though COPD prevalence in reported COVID-19 cases is low, COVID-19 infection is associated with significant severity and mortality in COPD. There was also increased risk of severe disease and mortality in current smokers. Effective preventive measures are urgently required to reduce COVID-19 risk on COPD patients and current smokers.

## Data Availability

Not applicable.

## Appendix

**Table S1:**
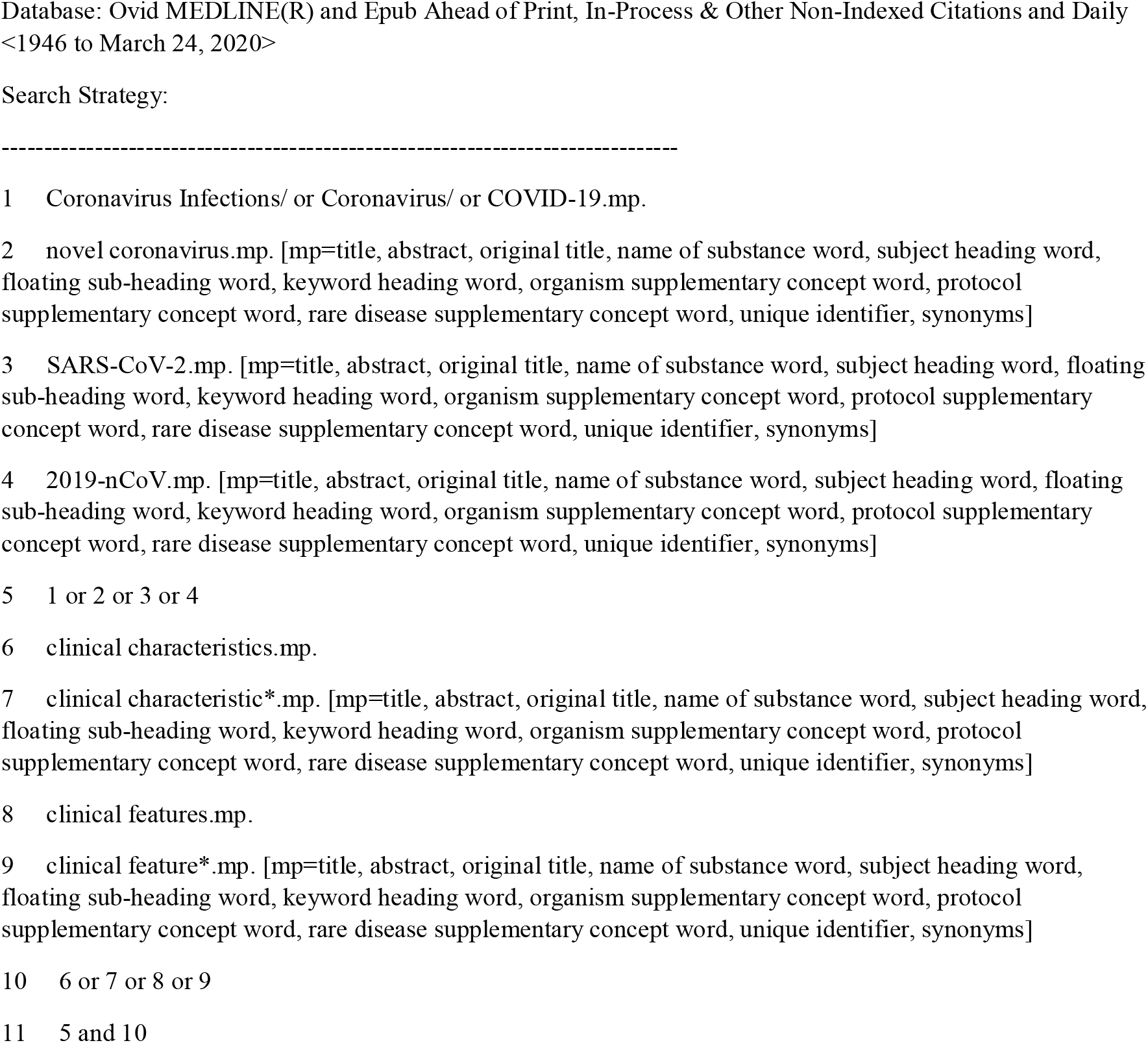
Medline search strategy.

**Table S2.**
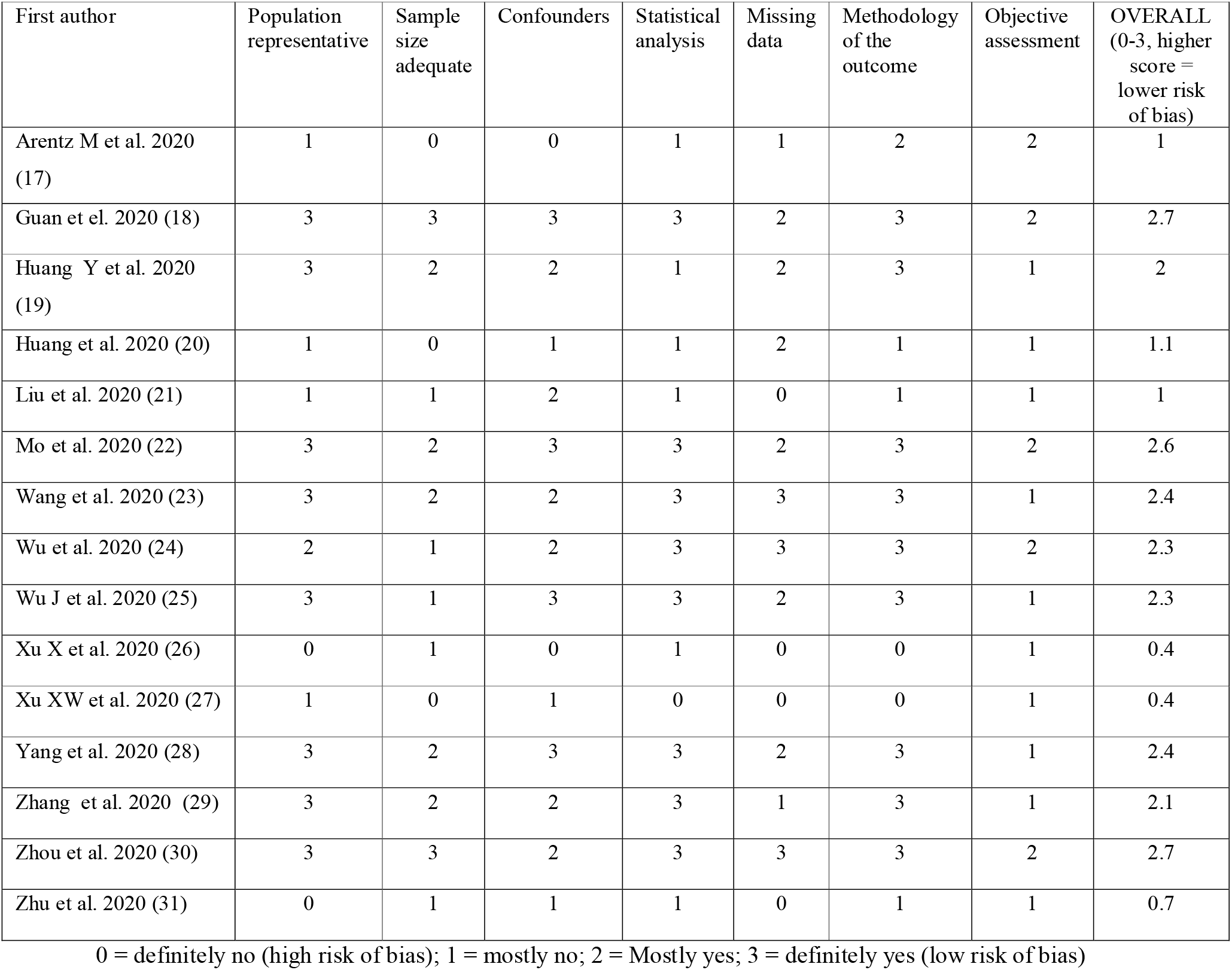
Quality assessment.

| First author | Population representative | Sample size adequate | Confounders | Statistical analysis | Missing data | Methodology of the outcome | Objective assessment | OVERALL (0-3, higher score = lower risk of bias) |
| --- | --- | --- | --- | --- | --- | --- | --- | --- |
| Arentz M et al. 2020 (17) | 1 | 0 | 0 | 1 | 1 | 2 | 2 | 1 |
| Guan et al. 2020 (18) | 3 | 3 | 3 | 3 | 2 | 3 | 2 | 2.7 |
| Huang Y et al. 2020 (19) | 3 | 2 | 2 | 1 | 2 | 3 | 1 | 2 |
| Huang et al. 2020 (20) | 1 | 0 | 1 | 1 | 2 | 1 | 1 | 1.1 |
| Liu et al. 2020 (21) | 1 | 1 | 2 | 1 | 0 | 1 | 1 | 1 |
| Mo et al. 2020 (22) | 3 | 2 | 3 | 3 | 2 | 3 | 2 | 2.6 |
| Wang et al. 2020 (23) | 3 | 2 | 2 | 3 | 3 | 3 | 1 | 2.4 |
| Wu et al. 2020 (24) | 2 | 1 | 2 | 3 | 3 | 3 | 2 | 2.3 |
| Wu J et al. 2020 (25) | 3 | 1 | 3 | 3 | 2 | 3 | 1 | 2.3 |
| Xu X et al. 2020 (26) | 0 | 1 | 0 | 1 | 0 | 0 | 1 | 0.4 |
| Xu XW et al. 2020 (27) | 1 | 0 | 1 | 0 | 0 | 0 | 1 | 0.4 |
| Yang et al. 2020 (28) | 3 | 2 | 3 | 3 | 2 | 3 | 1 | 2.4 |
| Zhang et al. 2020 (29) | 3 | 2 | 2 | 3 | 1 | 3 | 1 | 2.1 |
| Zhou et al. 2020 (30) | 3 | 3 | 2 | 3 | 3 | 3 | 2 | 2.7 |
| Zhu et al. 2020 (31) | 0 | 1 | 1 | 1 | 0 | 1 | 1 | 0.7 |

0 = definitely no (high risk of bias); 1 = mostly no; 2 = Mostly yes; 3 = definitely yes (low risk of bias)

## Notes

### Competing Interest Statement

The authors have declared no competing interest.

